# Prospective screening for monkeypox infection among users of HIV pre-exposure prophylaxis (PrEP) in Denmark

**DOI:** 10.1101/2023.06.23.23291799

**Authors:** Sofie Ellen Thomassen, Sebastian von Schreeb, Nikolai Søren Kirkby, Mette Pinholt, Anne-Mette Lebech, Gitte Kronborg, Frederik Neess Engsig

**Affiliations:** Department of Infectious Diseases, Copenhagen University Hospital - Amager and Hvidovre, Copenhagen Denmark; Infectious Disease Epidemiology and Prevention, Statens Serum Institut, Copenhagen, Denmark; Department of Clinical Microbiology, Copenhagen University Hospital - Rigshospitalet, Copenhagen, Denmark; Department of Clinical Microbiology, Copenhagen University Hospital - Amager and Hvidovre, Copenhagen, Denmark; Department of Infectious Diseases, Copenhagen University Hospital - Rigshospitalet, Copenhagen, Denmark; Faculty of Health and Medical Sciences, University of Copenhagen, Copenhagen, Denmark

## Abstract

**Introduction:** During the 2022 outbreak of mpox (previously called monkeypox), which primarily affected men who have sex with men (MSM), testing was mainly limited to individuals with symptoms of infection. Although sporadic cases of mpox continue to be diagnosed in Denmark, the benefit of screening asymptomatic high-risk populations, such as those using HIV pre-exposure prophylaxis (PrEP), is still unknown.

**Methods:** During the autumn of 2022, a rectal swab test for mpox PCR was included in the routine sexually transmitted infections (STI) screening for PrEP users.

**Results:** The screening included 224 asymptomatic men with a median age of 36.5 years. One patient (0.4%) tested positive for mpox. Ten (4.5%) and nine (4.0%) had chlamydia and gonorrhea, respectively.

**Discussion:** We found one case of mpox, who developed symptoms three days after testing positive. This finding may reflect low to non-existing levels of asymptomatic infection among mpox patients. Based on the current study, screening for asymptomatic mpox infection among PrEP users does not seem warranted.

## Introduction

Globally, 87.858 cases of mpox (previously called monkeypox) and 143 deaths have been confirmed since January 2022 and a vast increase in cases was reported from countries with no previous history of mpox transmission. Overall, 111 countries have reported cases, with Europe and the Americas being the most affected regions^1^. In Denmark, 196 cases have been confirmed since May 2022, with no confirmed cases since 26/01/2023. No deaths have been reported due to mpox in Denmark^2^. Most cases presented with mild disease symptoms, mainly rash and fever^1-4^.

In the 2022 mpox outbreak, a significant shift in transmission patterns has been observed, with the virus primarily spreading through sexual contact among men who have sex with men (MSM)^3^. Previously, human-to-human transmission was reported, but animal-to-human transmission, occurring via direct or indirect contact, was considered the predominant mode^5^.

Early studies from Central and Western African countries found no evidence of asymptomatic transmission and postulated that this mode of transmission had been ruled out^6^. However, during the current outbreak, it has been proposed that the possibility of asymptomatic transmission should be reconsidered^7^. If asymptomatic or subclinical infection is widespread, the current strategy of identification, isolation and contact tracing may be insufficient to contain mpox spread^8^. In Denmark as well as abroad, the use of HIV pre-exposure prophylaxis (PrEP) has been shown to be a risk factor for mpox infection^9,10^. For this reason, Danish authorities have offered smallpox vaccine (Imvanex®) to PrEP users since 12/08/2022, in accordance with World Health Organization (WHO) guidelines^11^. In the Danish PrEP program, participants are screened every three to six months for chlamydia, gonorrhea, HIV, syphilis as well as Hepatitis A, B (if not vaccinated) and C. Given that sporadic cases continued to be diagnosed in Denmark, we wanted to examine the prevalence of asymptomatic mpox infection among risk groups in order to evaluate if screening had a role to contain the mpox outbreak. If deemed beneficial, it may be practically feasible to include mpox in current PrEP screening programs. In this study, we set out to evaluate the incidence of asymptomatic mpox infection among PrEP users who were screened for sexual transmitted infections (STI) as part of the PrEP program.

## Methods

The study was performed at the Department of Infectious Diseases, Amager and Hvidovre Hospital and the Department of Infectious Diseases, Rigshospitalet, both part of Copenhagen University Hospital in the Capital Region of Denmark.

### Study population

The study enrolled PrEP users from the Danish PrEP Database who underwent routine STI screening during PrEP consultations or walk-in testing for STI, where mpox was not suspected, from 29/08/2022 to 13/10/2022.

### Laboratory analysis

Mpox, chlamydia and gonorrhea PCR testing was performed on rectal swaps. As testing capacity was limited, we chose to screen only from rectal mucosa as studies point towards high sensitivity at this location for mpox^10,12^.

At Department of Clinical Microbiology, Hvidovre, MPXV real-time PCR was performed using the Novaplex MPXV Assay (Seegene, Seoul, South Korea) on the CFX Real-Time PCR system (BIO-RAD, California, USA). Chlamydia (*Chlamydia Trachomatis)* and gonorrhea (*Neisseria Gonorrhoeae*) were tested using the Aptima Combo 2 assay on the Panther system (Hologic, San Diego, USA). At the Department of Clinical Microbiology, Rigshospitalet, q-PCR was performed using InfectionDetect, Mpox qPCR assay from PentaBase^13^. Test for chlamydia (*C. Trachomatis*) and gonorrhea (*N. Gonorrhoeae*) at the Department of Clinical Microbiology, Rigshospitalet was performed using Cobas, dual Swap samples, Roche Diagnostics.

The study was approved by the legal department of The Capital Region of Denmark (Authorization number P-2020-315).

## Results

A total of 224 male PrEP users were included in the study. This corresponds to 8% (83/1027) and 29% (141/479) of the PrEP users at Hvidovre Hospital and Rigshospitalet, respectively, included in The Danish PrEP Database per first of August 2022.

The study population were all MSM and had a median age of 36.5 years (interquartile range/IQR; 30-45).

### Mpox

All participants were tested once during the study period and four patients were tested twice on separate dates. One test (0.4%) was positive for mpox. The sample was taken from a man in his late forties in the beginning of September 2022. He showed no symptoms of mpox infection at the time of testing, reported no known exposure to mpox, had no recent travel history and reported that his partner had tested negative for mpox. The patient had received his first and only dose of smallpox vaccine 26 days prior to testing positive. Three days after testing positive, he reported three red lesions on his fingers and one lesion on his forehead but did not report having fever nor feeling unwell. No further contact with health authorities was made.

### Chlamydia and gonorrhea screening

A total of ten individuals (4.5%) tested positive for chlamydia and nine individuals (4.0%) tested positive for gonorrhea.

### Vaccine-status

At the time of mpox screening, 171 of the study population (76.3%) had received one dose of mpox vaccine and 78 (34.8%) had received second and last dose of vaccine. Among the 25 participants (11.2%) born before 1970 (assumed smallpox vaccinated as part of the previous Danish vaccination program^14^) 15 (6.7%) had received a booster vaccine against mpox.

A total of 64 participants (28.6%) were considered protected by the vaccine at the time of the mpox screening.

## Discussion

In this study we screened for asymptomatic mpox infection among PrEP users in routine testing for STI. During the study period, only one out of 224 participants (0.45%) tested positive for mpox, and he then presented symptoms of mpox three days later. Our findings indicate, that asymptomatic mpox infection is non-existing among PrEP users in Denmark.

Our result differs from those of the two other studies on the subject from Belgium and France. Ferré et al found a asymptomatic mpox prevalence of 4,9% in French MSMs on PrEP in June/July 2022 retrospectively and De Baetselier et al 1,8% in a population of MSM with/without HIV or PrEP in May 2022^15,16^. Several factors can explain this difference but besides differences in health care systems, our study was performed at a later stage of the outbreak where the incidence of mpox was lower and many persons at risk had received at least one mpox vaccination, which should confer some degree of protection^17^. The low incidence of mpox cases in Denmark at the time of screening could be related to an increased awareness in high-risk populations such as PrEP users and HIV infected, isolation and infection control strategies, testing of symptomatic individuals, contact tracing and vaccination of persons in high risk of mpox infection.

The patient who tested positive for mpox later developed mild symptoms of mpox infection and was thus pre-symptomatic at the time of testing. Pre-symptomatic patients with verified mpox infections have been detected in other screening studies and although viable mpox virus has been demonstrated in rectal swaps from apparently asymptomatic and non-vaccinated persons, the ability to transmit infection in this phase of the infection is uncertain ^10,15^. As such, the usefulness of diagnosing patients with mpox in the pre-symptomatic phase on mpox transmission is unknown.

Recent publications report 40-86% protection from mpox infection after 1-2 doses of Imvanex®/Jynneos® vaccine and apparently poor neutralizing capacity^17,18^. Our mpox infected patient only developed mild symptoms. Whether this is due to the size of inoculum, the fact that most mpox infected only develop mild symptoms or perhaps an effect of his single vaccination 26 days prior is unknown. Knowledge about viral transmission from asymptomatic and pre-symptomatic patients, vaccinated and un-vaccinated, still needs further research.

Our study shows that it is practically possible to include mpox in the STI screening program of PrEP users. The impact of screening PrEP users for asymptomatic mpox transmission can be evaluated from both medical and societal perspectives, considering costs such as resources, time, and money, as well as the potential physical and psychological effects on participants.

Screening from more anatomical sites could possibly increase our sensitivity and possibly the percentage of asymptomatic mpox patients. Still several studies point towards a high sensitivity at the rectal mucosa both during the viremic phase of illness and the symptomatic phase^10,12^. A major strength of the study is that we have complete follow-up on hospital contacts, STI testing and vaccination status. The incidence of chlamydia and gonorrhea confirmed that we screened a population with relevant risk behavior.

We evaluated the possible benefits of screening for asymptomatic mpox among PrEP users in Denmark during the global mpox outbreak in the fall 2022 and found only one case (0,45%). Our finding does not support screening for mpox among asymptomatic PrEP users.

## Data Availability

All data produced in the present study are available upon reasonable request to the authors

## Abbreviations

MSM: Men who have Sex with Men
HIV: Human Immunodeficiency Virus
PrEP: Pre-Exposure Prophylaxis
STI: Sexually Transmitted Infections
PCR: Polymerase Chain Reaction
WHO: World Health Organization

